# Board-Level Performance of Leading Open-Weight Vision-Language Models on the Japanese Diagnostic Radiology Board Examination: Reasoning, Image-Input, and Language Effects

**DOI:** 10.64898/2026.07.09.26357709

**Authors:** Yuki Sonoda, Yosuke Yamagishi, Yuichiro Hirano, Soichiro Miki, Takahiro Nakao, Shouhei Hanaoka, Yukihiro Nomura, Akiyoshi Hamada, Noriko Kanemaru, Rintaro Miyo, Masumi Mizuki Takahashi, Reina Hosoi, Takeharu Yoshikawa, Osamu Abe

## Abstract

**Purpose:** To evaluate the latest open-weight vision-language models (VLMs) on the Japanese Diagnostic Radiology Board Examination (JDRBE), assessing overall accuracy and the effects of image input, reasoning, and language.

**Materials and Methods:** In this retrospective study, 29 open-weight VLMs from 13 developers, released in or after January 2025, were evaluated on 327 image-bearing questions from four years of the JDRBE, a non-public benchmark with low risk of data leakage. Each question was answered by each model with and without the image(s), under three language conditions and with reasoning enabled and disabled. Accuracy was the primary outcome, and within-model differences were tested with paired bootstrap confidence intervals and sign-flip permutation tests with Benjamini–Hochberg correction.

**Results:** In the Japanese condition with image input and reasoning, the leading models reached 73.7% (gemma-4-31B-it), 73.1% (Qwen3.5-397B-A17B), and 72.1% (Kimi-K2.6). On the 2025 subset, these three models (74.1%–75.5%) scored above the mean accuracy of five newly board-certified radiologists who passed the 2025 examination (72%; range, 65%–83%). Accuracy broadly scaled with model size, although compact gemma-4-31B-it matched larger models. Enabling reasoning improved accuracy in nearly all models and the contribution of image input was larger when reasoning was enabled, particularly in higher-performing models. English prompts generally outperformed Japanese prompts.

**Conclusion:** Several open-weight VLMs, without medical adaptation, performed at or above the mean of newly board-certified radiologists on the JDRBE, with both model size and reasoning contributing. The highest Japanese-language accuracy came from a compact model suitable for parameter-efficient fine-tuning and serving on a single graphics processing unit.

**Summary Statement:** Several open-weight vision-language models, run on local infrastructure and without medical adaptation, performed at or above the mean of newly board-certified radiologists on the Japanese Diagnostic Radiology Board Examination.

**Key Points:**

- In this retrospective study of 29 open-weight vision-language models evaluated on 327 image-bearing questions from the Japanese Diagnostic Radiology Board Examination, the leading models reached 72.1%–73.7% accuracy in the Japanese condition; on the 2025 subset, three models (74.1%–75.5%) scored above the mean of five newly board-certified radiologists who passed that examination (72%).
- Enabling reasoning improved accuracy in nearly all models across language conditions (mean within-model gain, +4.5 to +8.0 percentage points), and significant contributions of image input were more frequent in the reasoning-enabled condition (22 of 56 instances vs 11 of 61 with reasoning disabled).
- The highest Japanese-language accuracy was achieved by gemma-4-31B-it (73.7%), a compact model that can be served on a single graphics processing unit and fine-tuned with parameter-efficient methods; it also used the fewest tokens among the leading models.

## Introduction

Language models are increasingly used in medicine and the life sciences, and extending them to interpret medical images alongside text would broaden their applications (1,2). Because medical data contain sensitive patient information, however, sending them to external cloud application programming interfaces (APIs) raises privacy and regulatory concerns (3). Handling image-bearing data while keeping it in-house therefore calls for open-weight models that run entirely on local infrastructure. Unlike closed models, open-weight models can be freely adapted for downstream applications, and they align with the transparency and auditability that regulators increasingly require of medical artificial intelligence (AI) (4,5).

Large language models (LLMs) have been shown to possess broad clinical knowledge (6), and their application to medicine, including radiology, is under active investigation (7). Early vision-language models (VLMs) interpreted medical images poorly: until around 2024, presenting the same questions with and without images did not improve scores across a range of radiology board examinations (8,9). This changed in 2025. In a recent study, Gemini 2.5 Pro, released in March 2025, was among the first models to benefit significantly from image input on a radiology board examination (10), and the later Gemini 3 Pro surpassed the highest-scoring of five newly board-certified radiologists on the same examination (11). Every model to reach these milestones, however, was a proprietary, closed API model.

Recent open-weight VLMs have grown markedly in size and capability and now approach closed models on general-purpose benchmarks (12). In medicine, open-weight models have so far been evaluated mostly on text-only examinations (13). Two open-weight VLMs were evaluated on a nuclear medicine board examination that included images, where both scored well below the proprietary models and image-containing questions were answered less accurately than text-only ones (14). Existing benchmarks, however, are of limited value for assessing how well open-weight VLMs interpret medical images. First, benchmarks published on the web may leak into training data, so that an evaluation can overstate a model’s true ability (15). Second, public medical visual-question-answering datasets such as VQA-RAD pose comparatively simple questions (16,17), and do not capture the integrative reasoning of routine practice, in which multiple imaging findings are combined to reach a differential diagnosis. Assessing the true ability of open-weight VLMs therefore calls for a benchmark that resists data leakage and demands the integrative interpretation of medical images.

We identified the Japanese Diagnostic Radiology Board Examination (JDRBE) as a benchmark that meets these needs. Candidates must complete at least five years of specialty training, and most questions require integrating imaging and clinical findings. The questions are restricted to members of the Japan Radiological Society and are not publicly available on the web, and no official answers are released.

Language models are known to perform better in English than in other languages, including Japanese (18). In multilingual medical examinations, an English-language system prompt has been reported to improve accuracy even when the question text was kept in the original language, while frontier models achieved comparable accuracy regardless of prompt language (19). How language conditions affect the accuracy of open-weight VLMs in medical image interpretation has not been examined.

We therefore aimed to evaluate the performance of the latest generation of open-weight VLMs on the JDRBE. Our main analyses assessed each model’s accuracy and the contribution of image input, measured by comparing performance with and without the image(s). We further examined the effect of reasoning, the effect of presenting questions entirely in English and of switching only the instruction language to English, and the number of tokens consumed in reasoning.

## Materials and Methods

This retrospective study used past JDRBE questions made available to members of the Japan Radiological Society. Because these materials contain no individually identifiable patient information, the institutional review board waived the requirement for approval.

### Dataset

We used, without modification, the dataset assembled by Miki et al. (11). It comprises only the image-bearing questions from the 2021, 2023, 2024, and 2025 JDRBE examinations. The reference answer for each question had been established by consensus among radiologists with 18, 23, and 30 years of experience, and some questions had more than one correct option. Each question presents five answer options. The 327 questions with confirmed answers formed the analysis set. Question texts averaged about 129 Japanese characters. For the EN-EN condition, each question text was translated with an LLM API (GPT-5.4) configured so that inputs and outputs were neither used for training nor retained. Radiologists with 3 and 4 years of experience independently reviewed every translation against the original, and six questions in which either reviewer identified a translation error were corrected.

### Model selection

We considered VLMs released on or after January 1, 2025 that accept image input, with at least 100,000 downloads in the preceding 30 days and at least 4 billion (4B) parameters, excluding models specialized for tasks such as translation and physical reasoning, as well as fine-tuned derivatives of other models. Within each developer, we excluded an earlier-generation family only when a newer generation offered a larger top model; otherwise, both generations were kept. These criteria were applied to a Hugging Face Hub API snapshot taken on May 17, 2026. As an exception, we replaced GLM-4.1V-9B-Thinking and GLM-4.5V with the newer GLM-4.6V and GLM-4.6V-Flash, although they had fewer than 100,000 downloads, to keep each developer represented by its latest generation.

All models were run in BF16 precision, with two exceptions: for Llama-4-Maverick-17B-128E-Instruct, BF16 inference was not feasible in our environment, so we used the official FP8 weights, a format generally reported to cause minimal accuracy loss (20), and Kimi-K2.6, a 1 trillion–parameter (1T) model released only in INT4, was run in that format. Two other models that did not run on our inference stack were excluded, yielding 29 models from 13 developers. The full list and the basis for each decision are detailed in Appendix S1.2.

### Inference and experimental conditions

All models were served on our own infrastructure and queried through an OpenAI-compatible API. Each question was evaluated both with image input (vision) and with the question text alone (text-only) to isolate the contribution of the image(s), under three language conditions (denoted instruction language–question text language) : a Japanese prompt with the Japanese question text (JA-JA), an English prompt with the translated question text (EN-EN), and an English prompt with the Japanese question text (EN-JA). The Japanese-prompt condition used the same prompt as Miki et al. (11). Models with a reasoning capability were tested with reasoning both enabled and disabled. Generation parameters followed each model’s recommended settings, with temperature set to 0 when no recommendation was found. Under these settings, models with a temperature above zero sampled stochastically and were run three times per condition, whereas those with temperature zero used greedy decoding and were run once. The full prompts and translation procedure are detailed in Appendix S1.1, and generation parameters and adjustments to the system role for each model are listed in Appendix S1.3.

### Language support and analysis populations

We classified each model’s Japanese support from the statements on vision-language support in its public model card or README. Models were grouped into three categories: Japanese explicitly listed as supported, no language specification, and supported languages listed but Japanese not included. For analyses that involved reading the Japanese question texts, namely the JA-JA and EN-JA conditions, we included the first two categories, comprising 18 models. The EN-EN condition included all 29 models. The classification and adoption status of each model are detailed in Appendix S1.4.

### Scoring

The final answer was automatically extracted from the end of the output, where the prompt instructed the models to state it, and compared with the reference answer. For questions with more than one correct option, a response was scored correct only if the set of selected options matched exactly. Outputs from which no answer could be extracted were scored incorrect.

### Statistical analysis

The primary outcome was accuracy. Accuracy was computed for each model under each condition. For models with stochastic generation, the mean of three runs served as the representative value. Each within-model difference was summarized as an effect size, defined as the per-question accuracy difference, averaged first over seeds and then across questions. The 95% confidence interval was obtained from a question-level paired bootstrap using the bias-corrected and accelerated method (21). Confidence intervals for the accuracy of a single model under a single condition were computed with the Wilson score method. Statistical significance was assessed by a two-sided sign-flip permutation test on the per-question differences (22) with Benjamini–Hochberg correction (23) at a false discovery rate below 0.05 within three families corresponding to the image, reasoning, and language comparisons, as detailed in Appendix S1.5. The comparison between the models and the radiologists was descriptive. Post hoc analyses examined the reasoning language, approximated per question by its dominant Unicode-based character class, and the associated token consumption, as detailed in Appendix S1.4. All analyses were performed with Python (version 3.12; Python Software Foundation).

## Results

We evaluated 29 models from 13 developers, each under all combinations of vision versus text-only, the three language conditions (JA-JA, EN-EN, EN-JA), and reasoning enabled versus disabled. The analysis set comprised 327 questions. The final answer was successfully extracted from 99% of outputs per model–condition pair on average (median, 100%). Extraction failures, caused mainly by format violations or repetition that exceeded the token limit, were concentrated in lower-performing models, and those outputs were scored incorrect (Appendix S1.5).

### Overall accuracy

Across all examination years, with image input and reasoning enabled, gemma-4-31B-it (73.7%), Qwen3.5-397B-A17B (73.1%), and Kimi-K2.6 (72.1%) achieved the highest accuracies in the Japanese condition. Under the EN-EN condition, Kimi-K2.6 achieved the highest accuracy (76.9%), followed by Qwen3.5-397B-A17B (76.0%). Under the EN-JA condition, with the Japanese question text retained but the instructions in English, Qwen3.5-397B-A17B achieved the highest accuracy (76.9%) (Table 1).

**Table 1.**
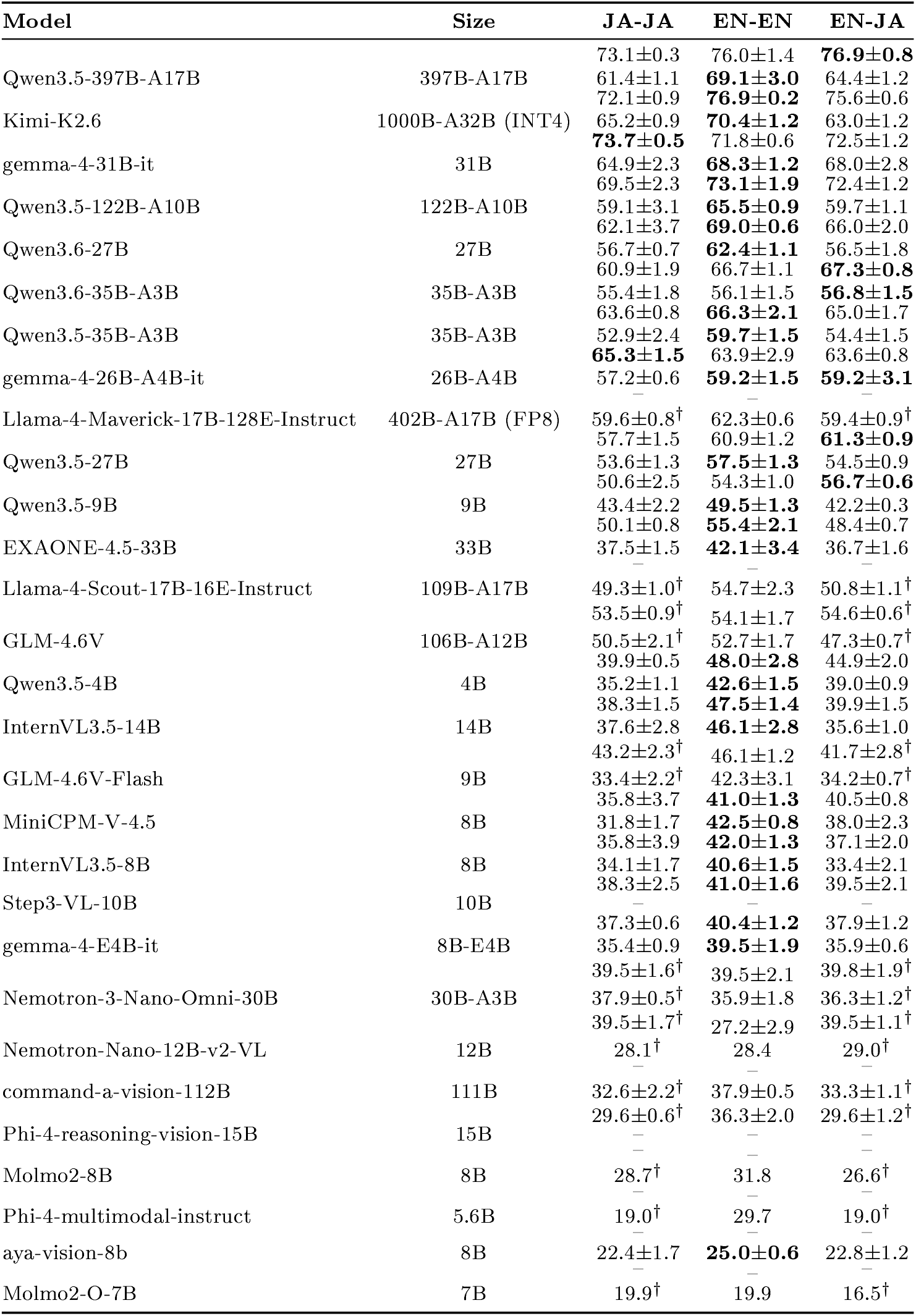
Accuracy by model, language condition, and reasoning mode. For each model, cells give accuracy (percent) for the Japanese (JA-JA), English (EN-EN), and English-instruction-with-Japanese-text (EN-JA) conditions, each cell showing accuracy with reasoning enabled (top line) and disabled (bottom line). A dash indicates an unavailable mode. For stochastic models these are the three-seed mean with standard deviation, whereas cells decoded deterministically (greedy, temperature 0) were run once and are shown without a standard deviation: all cells for Molmo2-8B, Molmo2-O-7B, and Phi-4-multimodal-instruct, and the reasoning-disabled cells for Nemotron-Nano-12B-v2-VL. Size gives total parameters and, for mixture-of-experts models, the active parameter count. Models are ordered by their highest accuracy across all conditions. A dagger (^*†*^) on a JA-JA or EN-JA cell marks a reference value from a model that does not officially support Japanese. For each Japanese-supported model, the highest accuracy across the three language conditions is shown in boldface, for reasoning enabled and disabled separately. E4B = Elective 4 billion parameters.

A previous study reported that five radiologists who passed the 2025 examination achieved a mean accuracy of 72% (standard deviation 7%, range 65–83%) on the 94 image-bearing questions (11). Because the examination was in Japanese, we compared model accuracy on the same 2025 subset under the corresponding Japanese condition. With reasoning enabled, the three leading models on this subset (Kimi-K2.6, 75.5%; gemma-4-31B-it, 74.5%; and Qwen3.5-397B-A17B, 74.1%) exceeded this mean, and the lower bounds of their 95% confidence intervals approached the lowest score among the five radiologists (Figure 1).

**Figure 1.**
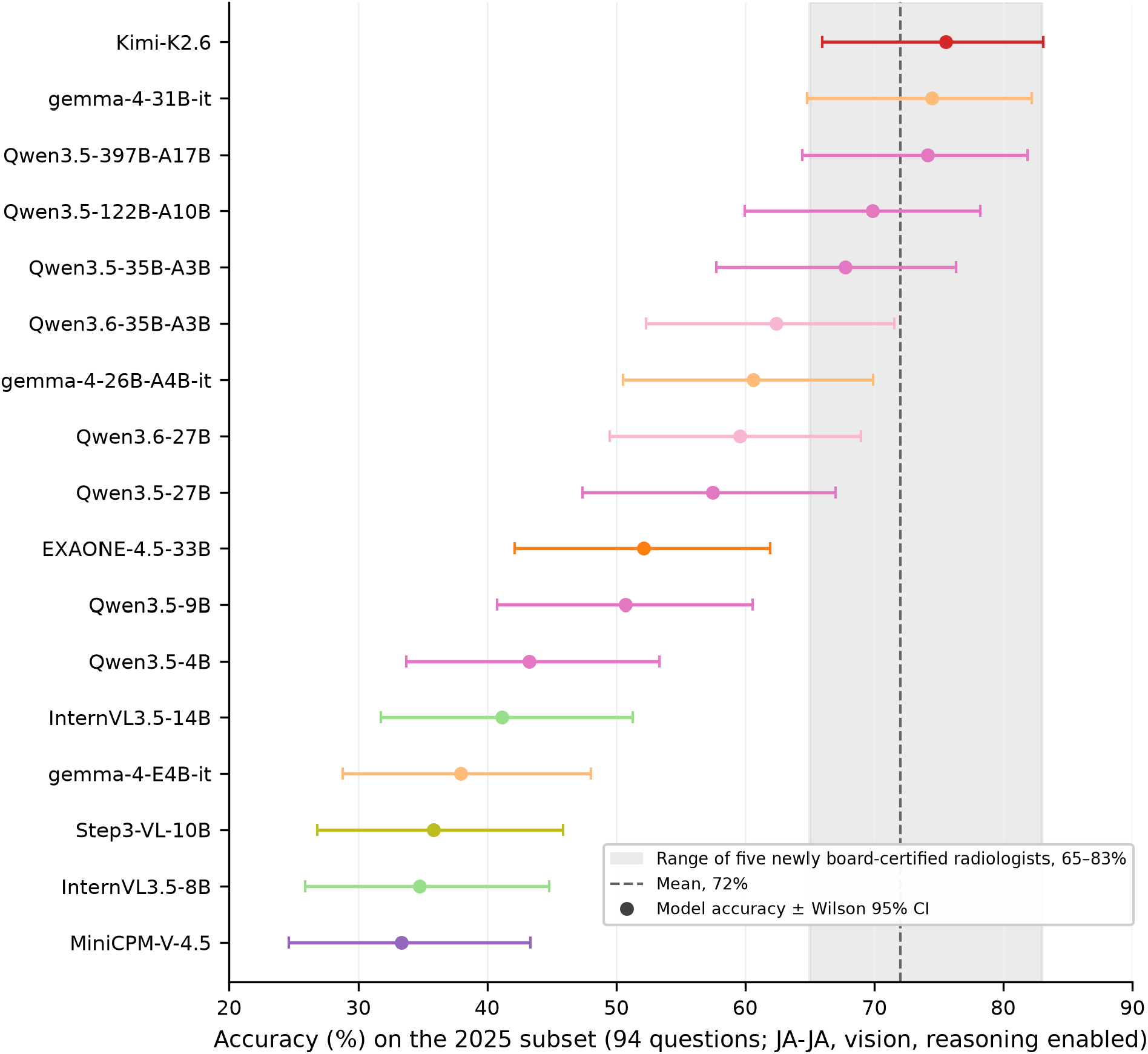
Accuracy on the 2025 examination subset relative to newly board-certified radiologists. Each point is the accuracy of one model on the 94 image-bearing questions of the 2025 administration, evaluated in the Japanese condition (JA-JA: Japanese instructions with Japanese question text) with image input (vision) and reasoning enabled and averaged over three seeds; horizontal bars give the Wilson 95% confidence interval (CI). The shaded band and dashed line mark the range (65% to 83%) and the mean (72%) of the five radiologists who passed the 2025 examination in a previous study (11). Models are ordered by accuracy. The 17 Japanese-supported models with a reasoning-enabled run on this subset are shown.

### Model size

Accuracy broadly reflected model size: larger models occupied the top of the ranking, and the smallest models fell well below them (Figure 2, Figure S1). However, performance did not scale uniformly across families, and gemma-4-31B-it, with 31B parameters, outperformed models an order of magnitude larger in the Japanese condition (Figure S1).

**Figure 2.**
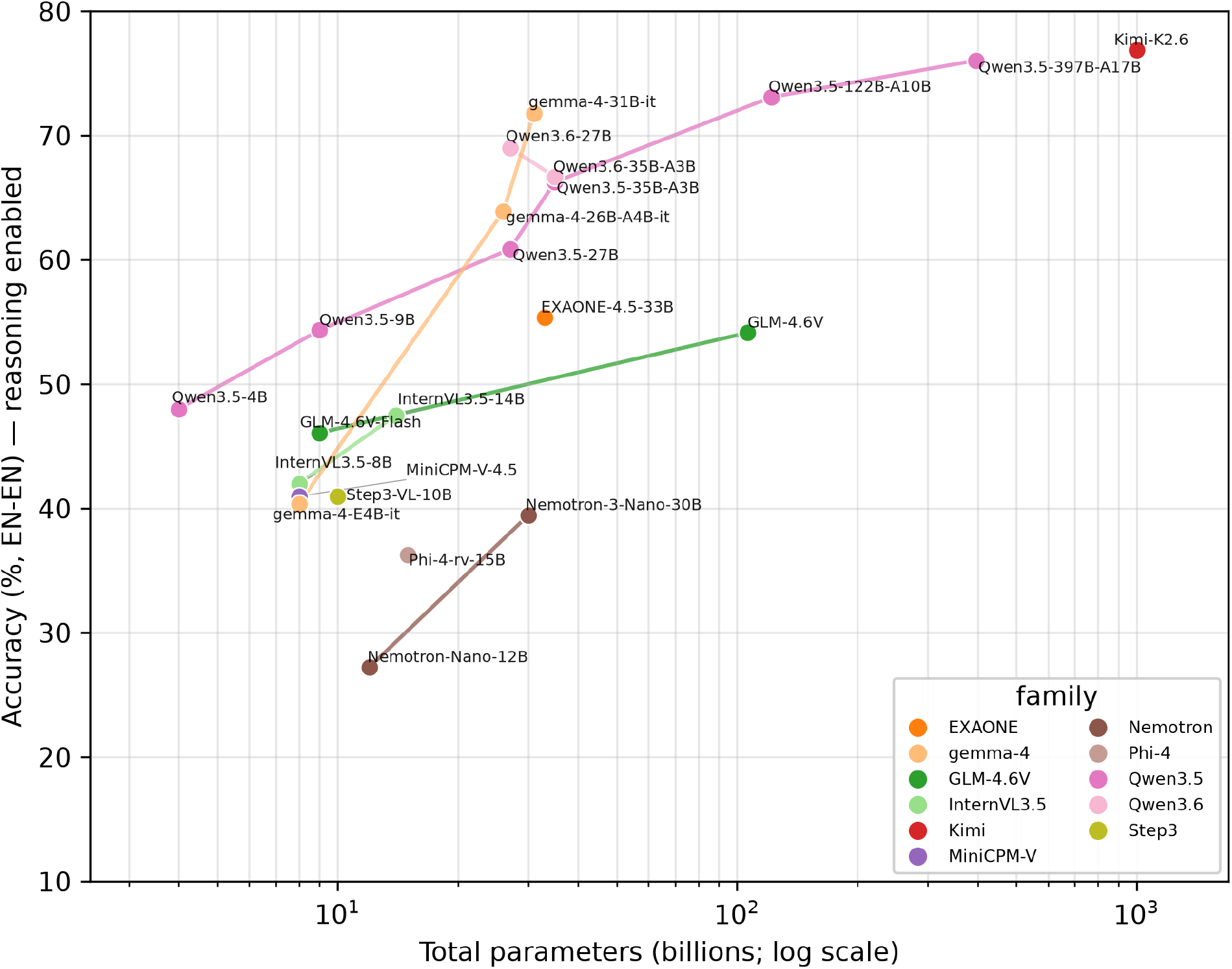
Accuracy versus model size in the fully English condition (EN-EN) with reasoning enabled, for all 22 models with a reasoning mode. Each point is a model plotted by total parameter count (logarithmic scale) against accuracy averaged over three seeds; lines join models of the same family and color denotes family.

### Effect of reasoning

Enabling reasoning improved accuracy in most models. Among models with both reasoning-enabled and reasoning-disabled modes, the mean within-model difference was positive in all three language conditions (+6.5 percentage points for JA-JA, +4.5 for EN-EN, and +8.0 for EN-JA), though the effect varied across models. The reasoning effect was positive in nearly all models in every language condition (16 of 16 for JA-JA, 18 of 20 for EN-EN, and 16 of 16 for EN-JA) and was significant in 13, 11, and 13 of these, respectively (Figure 3).

**Figure 3.**
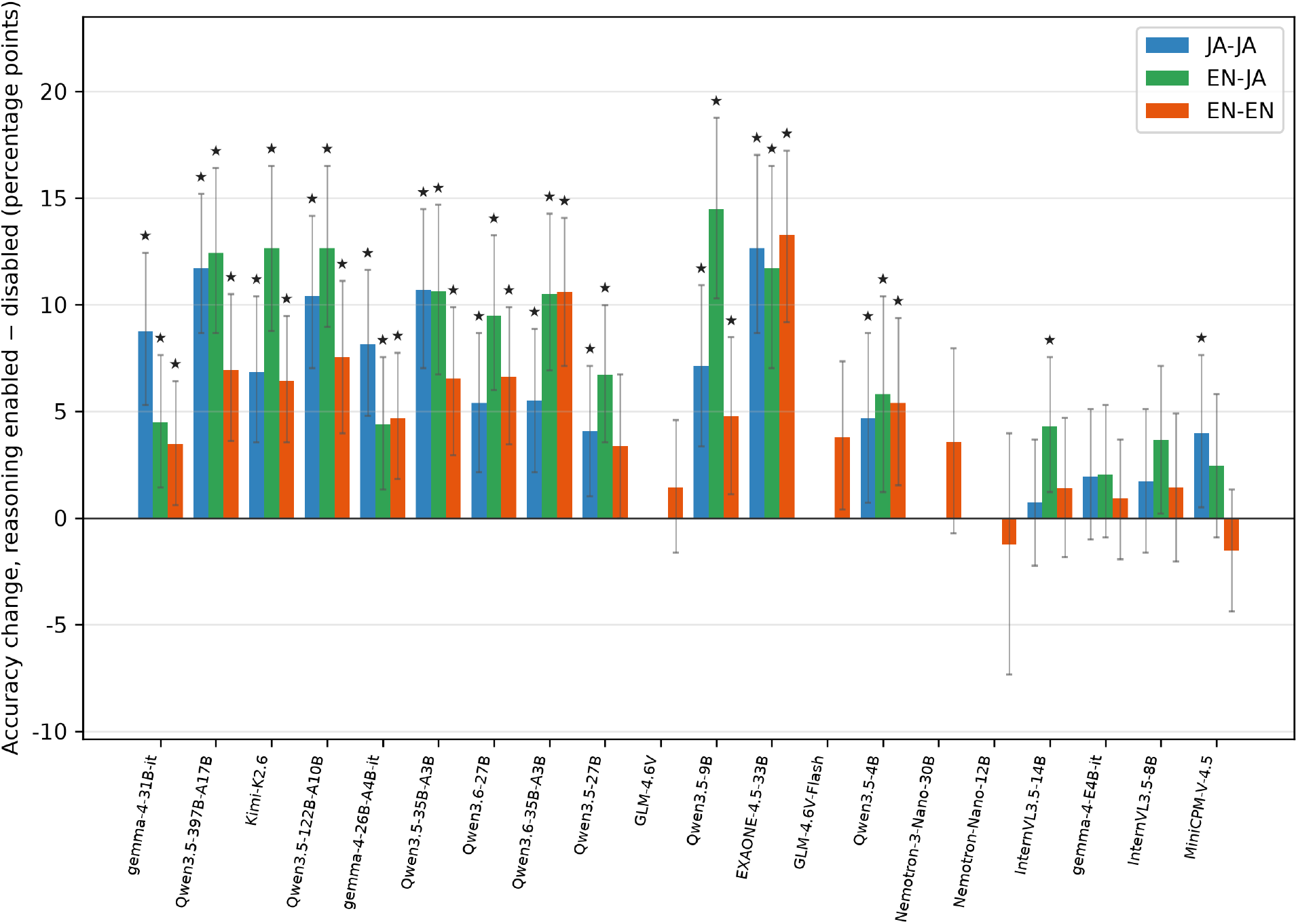
Elect of reasoning on accuracy by model and language condition, for the 20 models for which reasoning can be both enabled and disabled. For each model, bars give the change in accuracy from the non-reasoning to the reasoning mode (reasoning minus non-reasoning, in percentage points on scorable questions), averaged over three seeds, for the Japanese (JA-JA), English-instruction-with-Japanese-text (EN-JA), and fully English (EN-EN) conditions. Models that do not officially support Japanese are shown for the English condition only. Error bars are bias-corrected and accelerated 95% bootstrap confidence intervals. A star marks significance after Benjamini–Hochberg correction within the family (sign-flip permutation test, adjusted P *<* .05). Models are ordered by descending accuracy in the Japanese condition with reasoning enabled. Nemotron-3-Nano-30B = Nemotron-3-Nano-Omni-30B; Nemotron-Nano-12B = Nemotron-Nano-12B-v2-VL.

### Contribution of image input

The contribution of image input was quantified for each model as the difference in accuracy between the vision and text-only conditions (Figure 4). For some models this difference was near zero or negative. Significant positive contributions were more frequent with reasoning enabled and occurred mainly in the larger models. Across the three language conditions combined, image input significantly improved accuracy in 22 of 56 instances with reasoning enabled versus 11 of 61 with reasoning disabled (Figures 4, S2). A significant reduction occurred in two instances with reasoning enabled and one with reasoning disabled.

**Figure 4.**
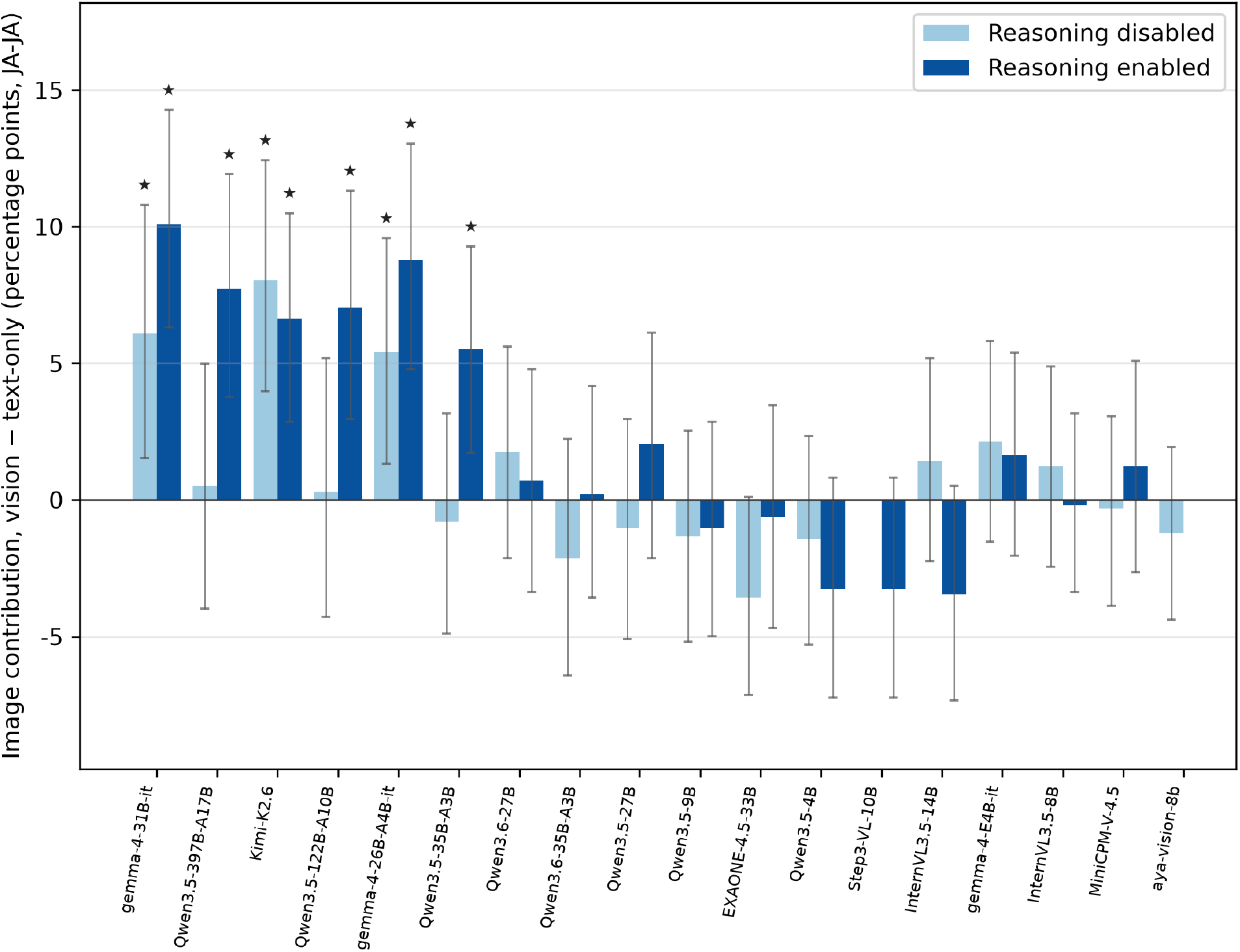
Contribution of image input by model in the Japanese condition (JA-JA), for the 18 Japanese-supported models. For each model, bars give the difference in accuracy between the vision and text-only settings (vision minus text-only, in percentage points on scorable questions), averaged over three seeds (single-run models are plotted as is), with reasoning disabled (light blue) and enabled (dark blue). Error bars, stars, and model order are as in Figure 3.

### Differences by language condition and token consumption

Conditions using English prompts generally outperformed the Japanese condition. With reasoning enabled, the differences were positive in most models for both EN-EN *−* JA-JA (15 of 17) and EN-JA *−* JA-JA (14 of 17) and were statistically significant in 7 of 17 and 7 of 17 models, respectively (Figure 5). With reasoning disabled, the differences were positive for EN-EN *−* JA-JA in all models (17 of 17) and for EN-JA *−* JA-JA in 11 of 17 models, with statistical significance observed in 10 of 17 and 1 of 17 models, respectively (Figure S3). The reasoning language varied across models: some reasoned in Japanese under JA-JA but switched to English under EN-JA, while others, including gemma-4-31B-it, reasoned in English regardless of the instruction language (Figure 6). Among the highest-performing models, Kimi-K2.6 and Qwen3.5-397B-A17B were among the models that switched, and their accuracy under EN-JA was significantly higher than under JA-JA, accompanied by an increase in reasoning-token consumption and a shift to English-language reasoning. Among the top-performing models, gemma-4-31B-it used the fewest reasoning tokens despite its high accuracy (Figure S4).

**Figure 5.**
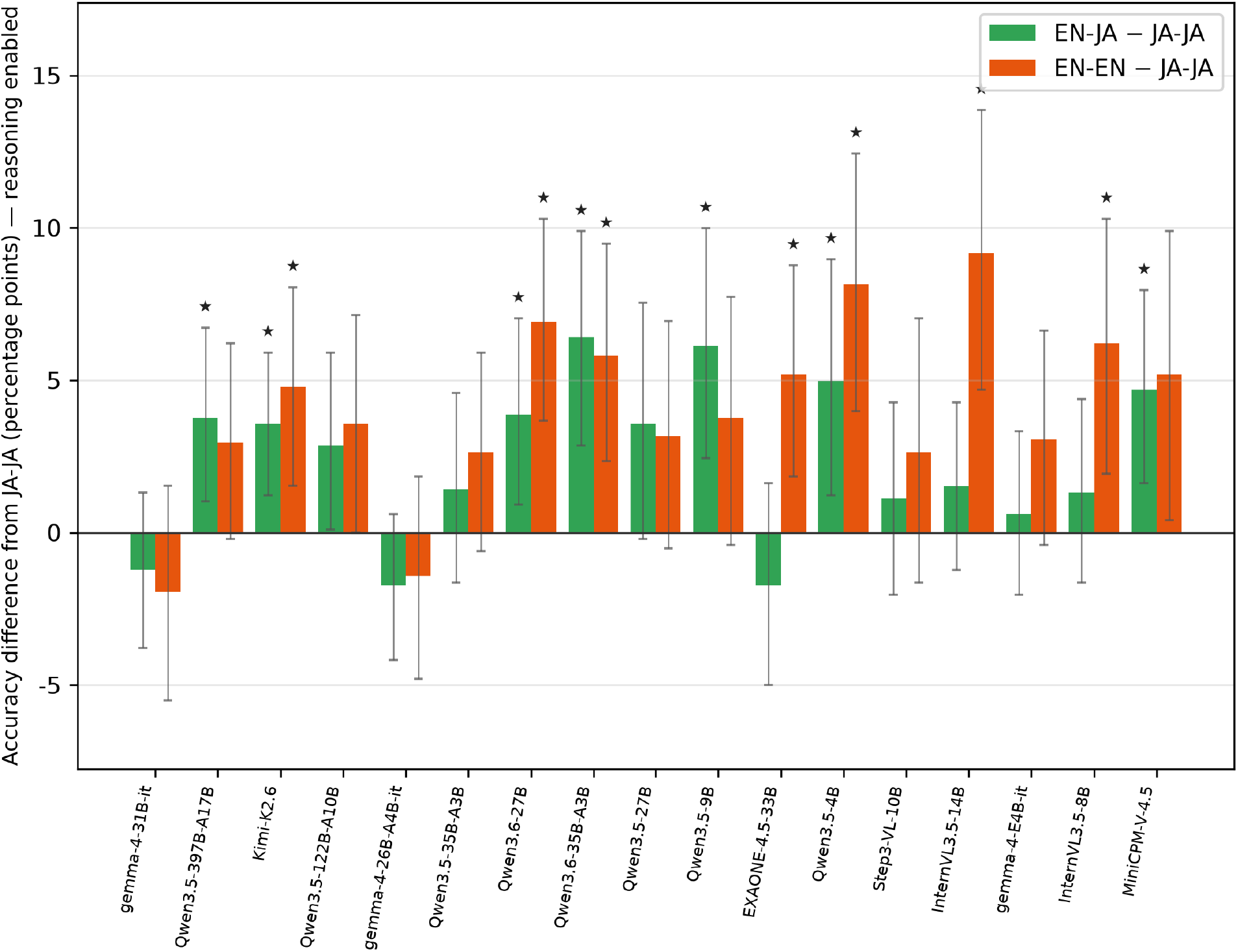
Effect of language condition relative to the Japanese baseline, with reasoning enabled, for the 17 Japanese-supported models evaluated under all three language conditions. For each model, bars give the change in accuracy from the Japanese condition for the English-instruction-with-Japanese-text condition (EN-JA minus JA-JA) and for the fully English condition (EN-EN minus JA-JA), in percentage points, averaged over three seeds. Positive values indicate higher accuracy than in Japanese. Error bars, stars, and model order are as in Figure 3.

**Figure 6.**
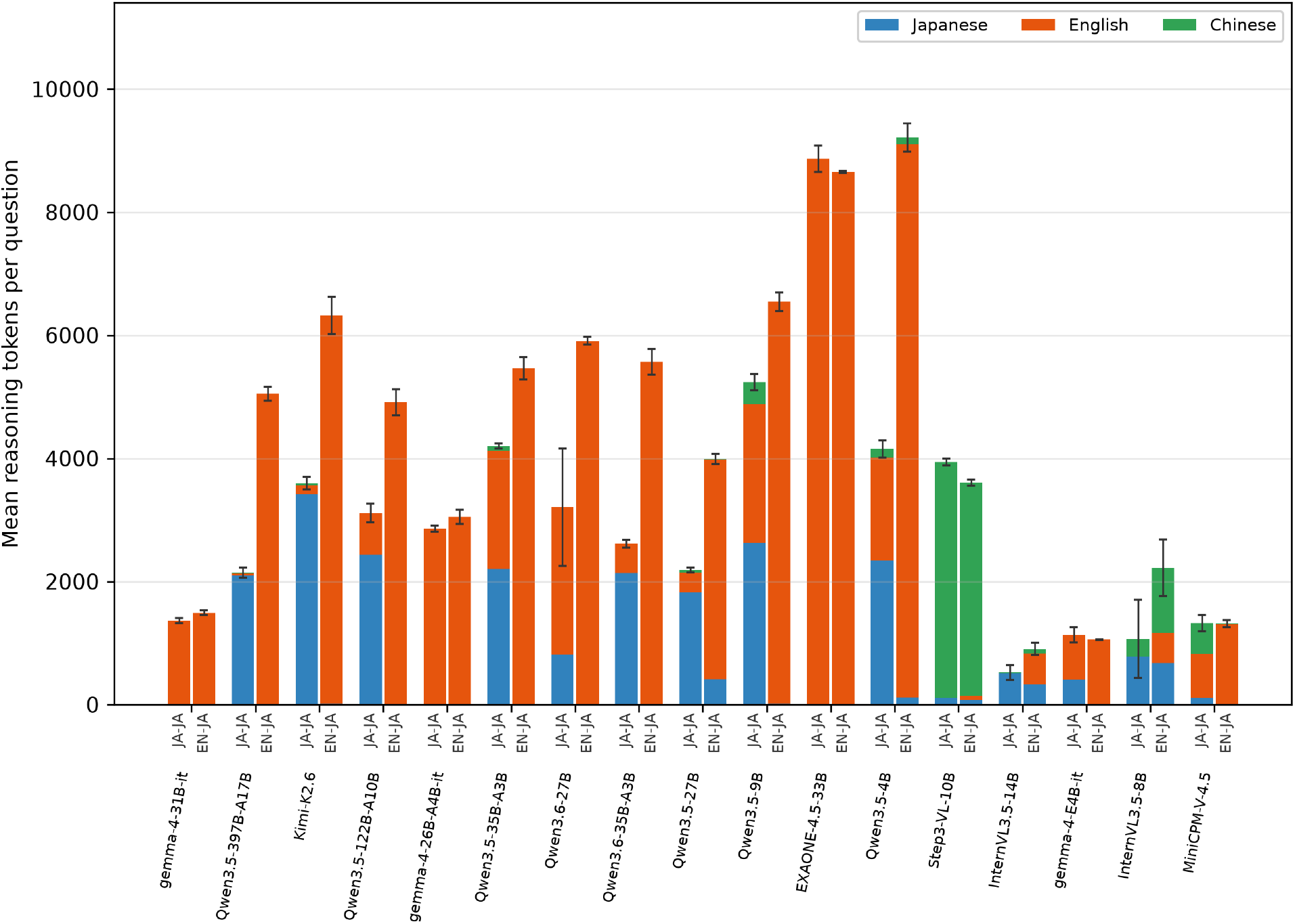
Reasoning-language composition and token consumption by model in the Japanese-text conditions, for the Japanese-supported models that have a reasoning mode. For each model, paired stacked bars give the mean number of reasoning tokens per question, partitioned by the dominant language of each question’s reasoning text (Japanese, English, or Chinese), determined approximately at the document level by Unicode-based character class (Appendix S1.4), under the Japanese condition (JA-JA) and the English-instruction-with-Japanese-text condition (EN-JA), with reasoning enabled and averaged over three seeds. Error bars are the standard deviation of the mean token count across the three seeds. Model order is as in Figure 3.

## Discussion

We evaluated 29 contemporary open-weight VLMs, spanning 4B to 1T parameters across 13 developers, on the JDRBE, a Japanese diagnostic radiology board examination with low risk of data leakage owing to its non-public questions. Smaller models fell well below board level, whereas the leading models surpassed the mean accuracy of the five radiologists who passed the 2025 examination (11). Enabling reasoning improved accuracy across language conditions and tended to increase the contribution of image input, particularly in the larger, higher-performing models.

Without any medical adaptation, the leading open-weight models surpassed the previous-generation Gemini 2.5 Pro (70.9%) in the Japanese condition and, under the EN-JA condition that retains the original Japanese questions, reached accuracies comparable to the current-generation closed models GPT-5.1 (74.6%) and Claude Opus 4.5 (76.8%), all evaluated on the same 327-question benchmark by Miki et al. (11). Unlike closed API models, whose responses may differ between calls and whose access can be withdrawn by providers or regulators (24–26), open-weight models have fixed weights that can be held on-site, offering reproducibility and permanence, which matter now that their accuracy approaches that of closed models and board-certified radiologists.

Within each model family, accuracy tended to increase with total parameter count, as reported in other domains and tasks (27,28). Across families, size alone did not determine accuracy. gemma-4-31B-it reached an accuracy comparable to that of models with hundreds of billions of parameters in the Japanese condition. Small enough for parameter-efficient fine-tuning and deployable on a single graphics processing unit (GPU; e.g., an NVIDIA RTX 6000 Pro Blackwell with 96 GB of memory), such a model may lower the barrier to medical AI research for groups working in non-English languages or with limited computational resources. Prior studies have already used open-weight models of this scale on local infrastructure for structured reporting and report de-identification (29,30).

Reasoning improved mean accuracy in every language condition, consistent with previous reports (31). Beyond the effect of reasoning, image input added further gains, mainly in the higher-performing models and more so when reasoning was enabled. This contrasts with findings from around 2024, when adding images did not improve, and at times reduced, commercial VLM accuracy on diagnostic-radiology board-examination questions (8,9).

The effect of language condition differed across models. For the two highest-scoring models, Qwen3.5-397B-A17B and Kimi-K2.6, accuracy under EN-JA was significantly higher than under JA-JA even though EN-JA retained the Japanese question text and switched only the instructions to English. Indeed, the highest accuracy observed in the entire study (76.9%; Qwen3.5-397B-A17B) was reached under EN-JA. In these models, the English instructions were accompanied by a shift of the reasoning language to English and an increase in token consumption, whereas gemma-4-31B-it, which reasoned in English regardless of the instruction language, showed no such gain. This pattern accords with reports that multilingual models reason internally in English (32) and answer more accurately when a task is routed through English (33), and with the finding that large reasoning models default to English-language reasoning and lose accuracy when reasoning is constrained to a lower-resource language (34). In practice, the instruction language may therefore be worth tuning, independently of the language of the clinical text.

This study has several limitations. First, the comparison with radiologists was descriptive, and we performed no formal statistical tests for superiority or equivalence because of the small sample sizes. However, the lower bounds of the leading models’ confidence intervals approached the lowest score among the five radiologists, suggesting that the proximity between the models and the radiologists is not an artifact of point estimation alone. Moreover, the benchmark comprises only the image-bearing subset of the full examination, which VLMs have been reported to answer less accurately than text-only questions (14), making the comparison conservative. Second, the contribution of image input was assessed at the population level through image ablation and does not address whether individual findings were correctly interpreted. Finally, although the JDRBE is an established standard for board certification, examination performance does not guarantee performance on real-world tasks beyond the examination.

In this study, leading open-weight VLMs, without medical adaptation, performed at or above the mean accuracy of newly board-certified radiologists on the JDRBE. Enabling reasoning improved accuracy in nearly all models, and image input contributed additional gains mainly in the higher-performing models with reasoning enabled, a capability that earlier-generation VLMs did not demonstrate on radiology board examinations. Switching only the instruction language to English, while retaining the Japanese question text, was sufficient to improve accuracy in some models, including those that scored at or above the mean of board-certified radiologists, although the magnitude and significance of this effect varied across models. Because open-weight models provide fixed, reproducible weights that can be hosted on-site and adapted to downstream tasks, and the top-performing model in the non-English condition can be served and fine-tuned on a single GPU, they may lower the barrier to medical AI research in resource-limited or non-English-speaking settings. Whether these results generalize to applied radiology tasks beyond board examinations remains to be established.

## Data availability

The code used for model inference and evaluation is available at https://github.com/miscelllaneous/jdrbe-vlm. The examination questions and the models’ outputs are not publicly available because the questions are copyrighted material of the Japan Radiological Society and the outputs are derived from them; these are available from the corresponding author on reasonable request.

## Acknowledgements

The Department of Computational Diagnostic Radiology and Preventive Medicine, The University of Tokyo Hospital, is sponsored by HIMEDIC Inc. and Siemens Healthcare.

## Author contributions

Y.S.: Study concept and design, software development, running the experiments, data collection, formal analysis, visualization, and drafting of the manuscript.

Y.Y.: Data curation, investigation, and validation.

Y.H.: Software development and data collection.

S.H., S.M., T.Y.: Preparation of the reference answers.

A.H., N.K., R.M., M.M.T., R.H.: Preparation of the answer data.

S.H., S.M., T.Y., T.N., Y.N., O.A.: Validation.

All authors critically revised the manuscript for important intellectual content, approved the final version, and agree to be accountable for all aspects of the work.

