## supplement for "Board-Level Performance of Leading Open-Weight Vision-Language Models on the Japanese Diagnostic Radiology Board Examination: Reasoning, Image-Input, and Language Effects"

### Appendix S1. Detailed Methods

#### S1.1 Prompts and language conditions

Three language conditions were defined by the language of the instruction (system prompt) and of the question text (denoted instruction language–question text language): JA-JA (Japanese instruction, Japanese question text), EN-EN (English instruction, English-translated question text), and EN-JA (English instruction, Japanese question text). For the image ablation, every question was presented twice, once with its image(s) and once with the text alone. The text-only variants replaced the sentence introducing the question with one noting that only the question text is provided. The with-image prompts for each condition are reproduced below, followed by the EN-EN text-only prompt as a representative example.

**JA-JA, with image:**

> あなたはこれから放射線科診断専門医試験を受験する放射線科医です。以下に専門医試験の問題を提示しますので、解答をお願いします。また、解答に至った思考過程も簡潔に説明してください。自信がない場合でも必ず解答を選択して答えることが強制されます。最終行に "ANSWER: x" または "ANSWER: x, y" の形で選択した選択肢を明示してください。

**EN-EN, with image:**

> You are a radiologist taking a radiology board certification exam. A board exam question is presented below. Please provide your answer and briefly explain your reasoning process. You must select an answer even if you are not confident. On the final line, state your selected option(s) as "ANSWER: x" or "ANSWER: x, y".

**EN-JA, with image:**

> You are a radiologist taking a radiology board certification exam. The question below is written in Japanese. Please provide your answer and briefly explain your reasoning process. You must select an answer even if you are not confident. On the final line, state your selected option(s) as "ANSWER: x" or "ANSWER: x, y".

**EN-EN, text only (no image):**

> You are a radiologist taking a radiology board certification exam. Note that only the question text is provided; no images are given. Please provide your answer and briefly explain your reasoning process. You must select an answer even if you are not confident. On the final line, state your selected option(s) as "ANSWER: x" or "ANSWER: x, y".

**Translation prompt (English conditions). The system prompt used to translate each Japanese question text into English was:**

> You are a professional translator specializing in diagnostic radiology. Translate the given Japanese radiology specialist exam question into English. Preserve the original formatting exactly: keep option labels (a, b, c, d, e) and line breaks. Do not add explanations or notes. Output only the translation.

### S1.2 Model selection

Open-weight VLMs were enumerated from the Hugging Face Hub by 30-day download count. At the level of each model size, a candidate entered the baseline pool only if it was a general-purpose VLM accepting image input (pipeline image-text-to-text or any-to-any), released on or after 2025-01-01, with at least 100,000 30-day downloads and an effective or advertised size of at least 4B. Quantized copies, base-weight checkpoints, duplicate mirrors, and reasoning-mode/instruct variants of the same checkpoint were collapsed to a single model, yielding a baseline pool of 54 models. These criteria were applied to a Hugging Face Hub API snapshot taken on May 17, 2026. From this pool models were removed in four steps, leaving 29 (Figure S5). First, two special-purpose models were removed: the translation-specialized TranslateGemma-4B and the physical-reasoning Cosmos-Reason2-8B.

Second, four fine-tuned derivatives of another released model were removed. By a derivative we mean a model obtained by further post-training a third party's released model and distributed as a variant, namely the Qwen-derived Bee-8B-RL and R-4B, the Llama-3.1-derived Llama-3.1-Nemotron-Nano-VL-8B, and the Gemma-derived MedGemma-1.5-4B.

Third, seventeen older-generation models that were superseded within their own developer were removed. Supersession was resolved across modality variants and judged by maximum parameter count, so that an older generation was dropped when the same developer offered a newer generation of at least equal maximum size. On this basis we removed, from Google, Gemma-3-12B, Gemma-3-4B, and Gemma-3-27B; from Alibaba/Qwen, Qwen3-Omni-30B, Qwen2.5-Omni-7B, Qwen3-VL-8B, Qwen3-VL-4B, Qwen3-VL-235B-A22B, Qwen3-VL-32B, Qwen3-VL-30B-A3B, Qwen2.5-VL-7B, Qwen2.5-VL-72B, and Qwen2.5-VL-32B, all superseded by the Qwen3.5 and Qwen3.6 families, which were themselves both retained because neither exceeded the other's maximum size; from NVIDIA, Eagle2.5-8B, superseded by the newer and larger Nemotron vision-language families; from OpenGVLab, InternVL3-8B and InternVL3-14B, superseded by InternVL3.5; and from Moonshot AI, Kimi-K2.5, superseded by Kimi-K2.6. In addition, GLM-4.1V-9B-Thinking and GLM-4.5V from Z.ai were replaced by GLM-4.6V and GLM-4.6V-Flash to keep each developer represented by its newest generation; as an exception, neither replacement model met the download threshold.

Fourth, two models that met all criteria but did not run on our inference stack were removed: LLaVA-OneVision-1.5-8B and StepFun Step3.

This procedure yielded 29 models from 13 developers.

### S1.3 Generation parameters and inference handling

Models were served on NVIDIA GH200 Grace Hopper nodes (pipeline parallelism across up to 12 nodes for the largest models), except Phi-4-multimodal-instruct, Phi-4-reasoning-vision-15B,

Nemotron-3-Nano-Omni-30B, and Nemotron-Nano-12B-v2-VL, which were served on a single NVIDIA RTX 6000 Pro Blackwell Max-Q workstation GPU (96 GB VRAM). The inference engine, library versions, GPU configuration, and generation parameters per model are given in Table S1. Generation parameters were taken from each model's official model card, README, or Hugging Face generation_config.json. For models without a recommended configuration, temperature was set to 0. The maximum number of output tokens was set for each model to 40,000 tokens or, if smaller, to the model's serving context window (vLLM/SGLang –max-model-len, i.e. the maximum input+output sequence length allowed by the serving instance) minus 4,096 tokens.

Reasoning was toggled via the chat-template parameter enable_thinking or an equivalent model-specific method documented in each model card. Phi-4-reasoning-vision-15B always reasons and was run in the reasoning-enabled condition only. Step3-VL-10B likewise lacks a reasoning-disabled mode and was run in the reasoning-enabled condition only, whereas aya-vision-8b has no reasoning mode and was run in the reasoning-disabled condition only. For models whose chat template does not accept a separate system role (Phi-4-reasoning-vision-15B, Molmo2-8B, Molmo2-O-7B, aya-vision-8b), the system prompt was concatenated into the user message.

#### S1.4 Analysis populations

Japanese-language support for each model was classified from its public model card or README into three categories:

Japanese explicitly listed as supported (2 models): aya-vision-8b, EXAONE-4.5-33B.

No language specification in model card (16 models): gemma-4-26B-A4B-it, gemma-4-31B-it, gemma-4-E4B-it, InternVL3.5-8B, InternVL3.5-14B, Kimi-K2.6, MiniCPM-V-4.5, Qwen3.5-4B, Qwen3.5-9B, Qwen3.5-27B, Qwen3.5-35B-A3B, Qwen3.5-122B-A10B, Qwen3.5-397B-A17B, Qwen3.6-27B, Qwen3.6-35B-A3B, Step3-VL-10B.

Supported languages listed in model card, Japanese not included (11 models): command-a-vision-112B, GLM-4.6V, GLM-4.6V-Flash, Llama-4-Maverick-17B-128E-Instruct, Llama-4-Scout-17B-16E-Instruct, Molmo2-8B, Molmo2-O-7B, Nemotron-3-Nano-Omni-30B, Nemotron-Nano-12B-v2-VL, Phi-4-multimodal-instruct, Phi-4-reasoning-vision-15B.

The first two categories (18 models total) were designated as adopted models (referred to as Japanese-supported models in the figure legends) for the JA-JA and EN-JA analyses. The EN-EN condition (English-translated question text) included all 29 models. Language-condition comparisons were restricted to the 17 adopted models with a reasoning mode.

Table S1 lists all 29 models with their developer, reasoning-mode availability, and Japanese-language support classification.

The dominant language of each question's reasoning output was approximated by Unicode-based character class. For each output, characters were counted in three categories: kana (hiragana, katakana, and half-width katakana), CJK ideographs, and Latin letters. Fractions were computed over the sum of these three categories. An output was labeled Japanese if the kana fraction was at least 5% and the combined kana-and-CJK fraction was at least 30%, Chinese if it was not Japanese and the CJK fraction was at least 30%, English if it was neither and the Latin fraction was at least 50%, and otherwise other.

#### S1.5 Software and statistics

Accuracy was computed for each model under each condition; for models with stochastic generation the mean of three runs served as the representative value, with variability summarized by the standard deviation and coefficient of variation. The contribution of image input, the effect of reasoning, and the difference between Japanese and English input were each quantified as a within-model difference in accuracy.

Each within-model difference (image contribution, reasoning effect, language effect) was summarized by an effect size: the mean across questions of the per-question accuracy difference, where each question's difference was first averaged over its available seeds. A 95% confidence interval was obtained from a question-level paired bootstrap (10,000 resamples of the questions with replacement) using the bias-corrected and accelerated (BCa) method. A two-sided p-value was obtained by a sign-flip permutation test on the per-question differences (1,000,000 sign assignments). Multiplicity was controlled with the Benjamini–Hochberg procedure (false discovery rate $< 0.05$) within three pre-specified families corresponding to the three comparison axes, namely image contribution (117 tests: vision versus text-only for each model in each of the three language conditions and two reasoning modes), reasoning effect (52 tests: reasoning enabled versus disabled for each model in each language condition), and language effect (68 tests: the EN-EN − JA-JA and EN-JA − JA-JA contrasts for each model in each reasoning mode), 237 tests in total.

Tokens in each output were counted with the model's own tokenizer. Analyses were run under Python 3.12 with numpy 2.4, scipy 1.13, pandas 2.2, and transformers 4.40 or later.

A valid answer could be parsed from at least 90% of responses in nearly all model–condition pairs (median, 100%). The only pairs falling below 90%, all in conditions with English instructions, were Nemotron-Nano-12B-v2-VL (EN-EN reasoning, 68.3%; EN-EN without reasoning, 77.7%), Nemotron-3-Nano-Omni-30B (EN-EN reasoning, 86.4%), and Molmo2-O-7B (EN-EN without reasoning, 89.6%; EN-JA without reasoning, 89.0%). Full per-model extraction rates are provided in the Supplementary Data.

### Supplementary Figures

### Supplementary Table

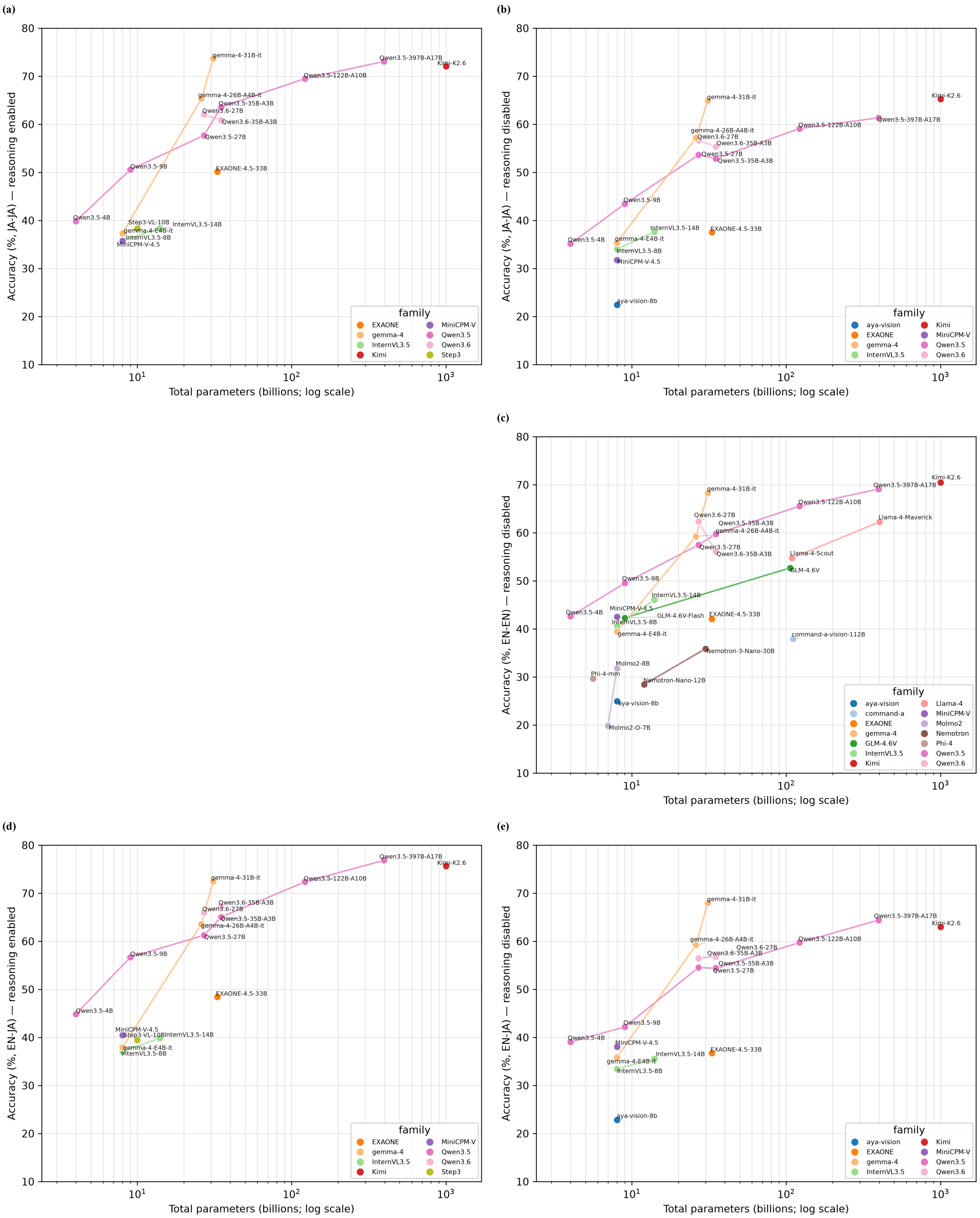


**Figure S1:** Accuracy versus model size for the five language-and-reasoning conditions not shown in Figure 2. Each point is a model plotted by total parameter count (logarithmic scale) against accuracy averaged over three seeds (single-run models are plotted as is); lines join models within a family. (a) Japanese condition, reasoning enabled (JA-JA, 17 models). (b) Japanese condition, reasoning disabled (JA-JA, 17 models). (c) Fully English condition, reasoning disabled (EN-EN, 27 models). (d) English-instruction-with-Japanese-text condition, reasoning enabled (EN-JA, 17 models). (e) English-instruction-with-Japanese-text condition, reasoning disabled (EN-JA, 17 models).

(a)

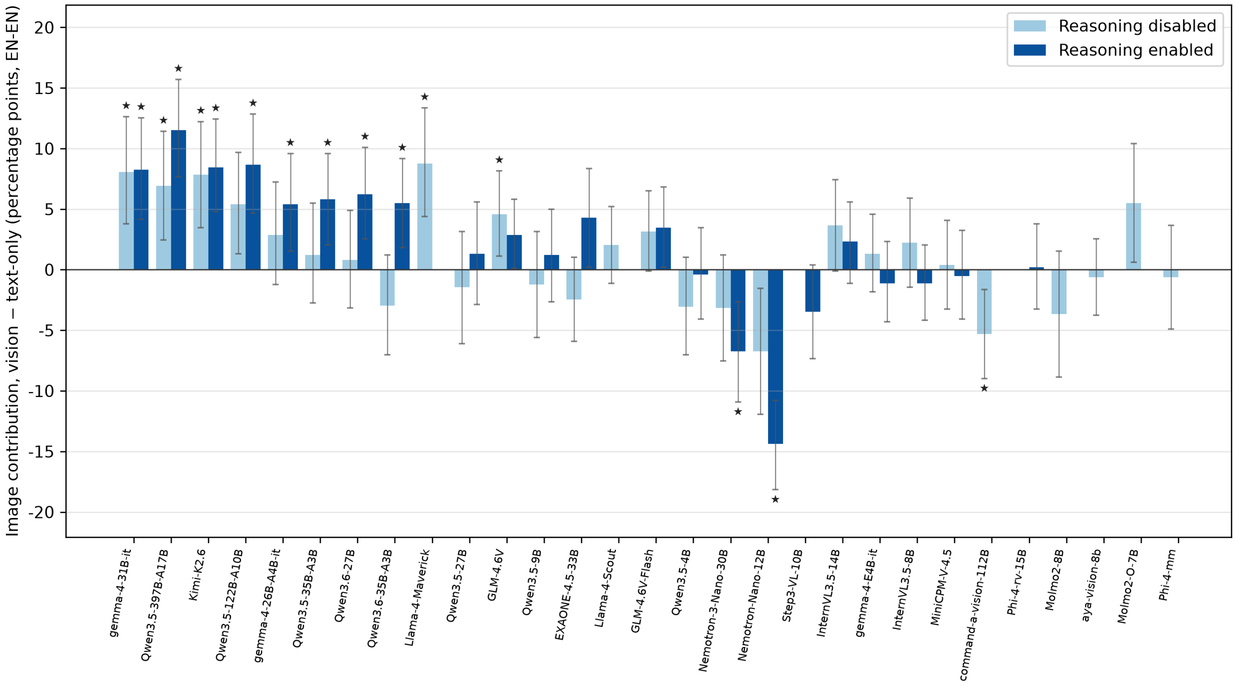


(b)

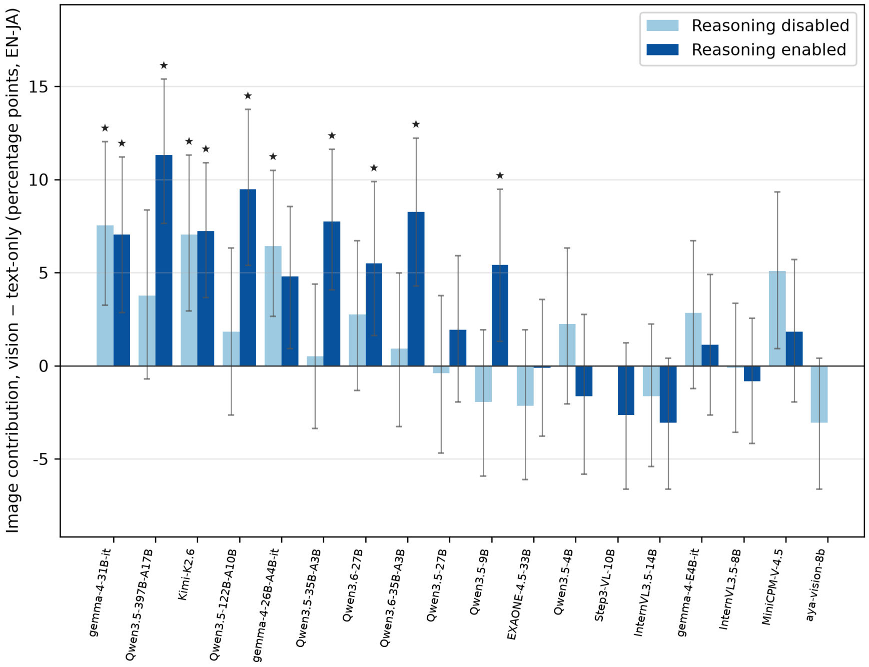


**Figure S2:** Contribution of image input by model in (a) the fully English condition (EN-EN) and (b) the English-instruction-with-Japanese-text condition (EN-JA). As in Figure 4, for each model the bars give the change in accuracy when images were provided relative to the text-only setting (vision minus text, in percentage points on scorable questions), averaged over three seeds (single-run models are plotted as is), with reasoning disabled (light blue) and enabled (dark blue). Error bars and stars are as in Figure 3. Models are ordered by descending accuracy in the Japanese condition. Panel (a) shows all 29 models; panel (b) is restricted to the 18 Japanese-supported models. Abbreviations on the horizontal axis: Llama-4-Maverick = Llama-4-Maverick-17B-128E-Instruct; Llama-4-Scout = Llama-4-Scout-17B-16E-Instruct; Nemotron-3-Nano-30B = Nemotron-3-Nano-Omni-30B; Nemotron-Nano-12B = Nemotron-Nano-12B-v2-VL; Phi-4-mm = Phi-4-multimodal-instruct; Phi-4-rv-15B = Phi-4-reasoning-vision-15B.

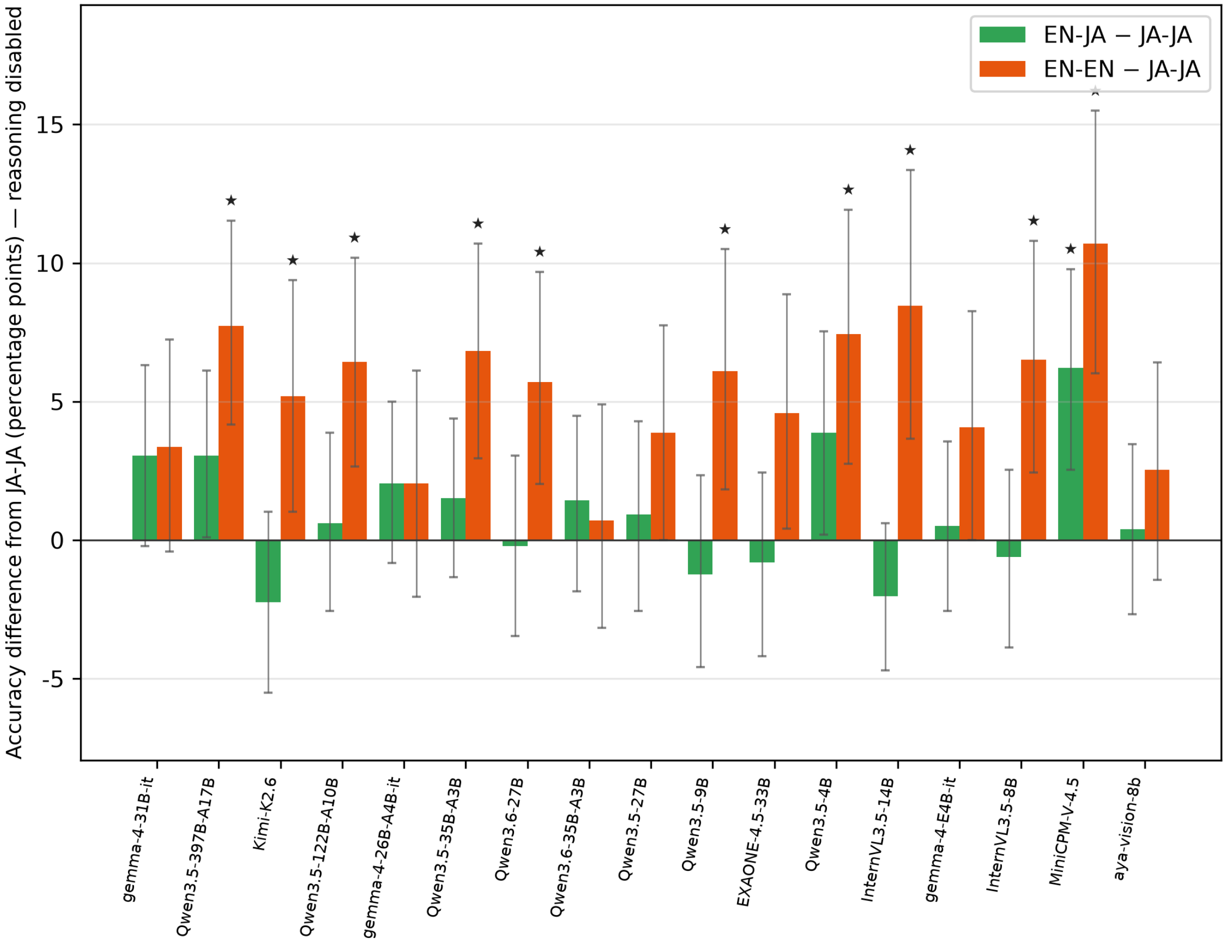


**Figure S3:** Effect of language condition relative to the Japanese baseline with reasoning disabled. As in Figure 5, bars give the change in accuracy from the Japanese condition for the EN-JA condition (EN-JA minus JA-JA) and the fully English condition (EN-EN minus JA-JA), in percentage points, averaged over three seeds. Error bars and stars are as in Figure 3. Restricted to the 17 Japanese-supported models evaluated under all three language conditions with reasoning disabled.

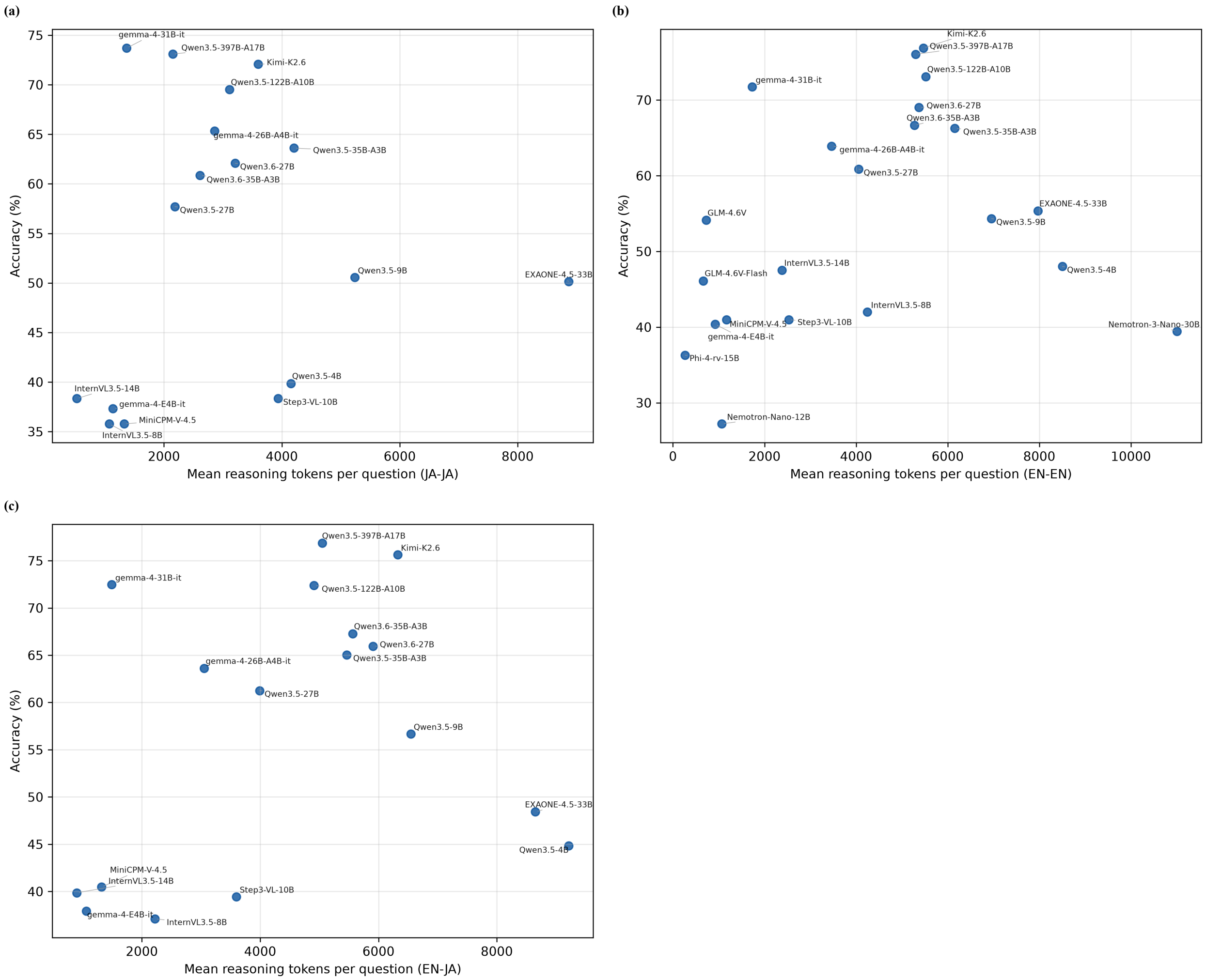


**Figure S4:** Reasoning-token consumption versus accuracy for each model with reasoning enabled. Each point is a model plotted by the mean number of reasoning tokens per question against accuracy averaged over three seeds. (a) Japanese condition (JA-JA, 17 models). (b) Fully English condition (EN-EN, 22 models). (c) English-instruction-with-Japanese-text condition (EN-JA, 17 models).

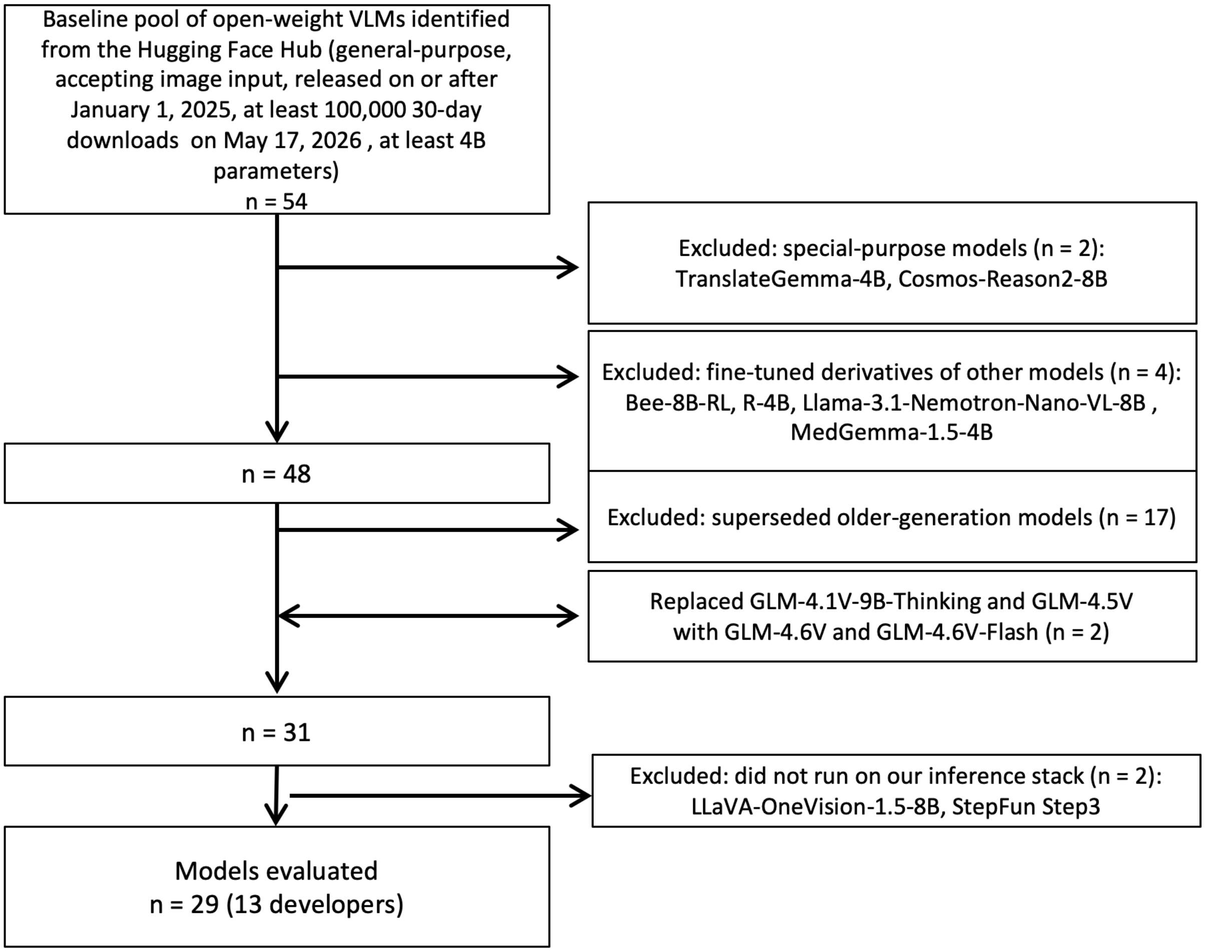


**Figure S5:** Model-selection flow diagram. From a baseline pool of 54 open-weight VLMs, models were excluded in four steps: special-purpose models (n = 2), fine-tuned derivatives (n = 4), superseded older-generation models (n = 17), and models that did not run on the inference stack (n = 2), yielding 29 evaluated models. In addition, GLM-4.1V-9B-Thinking and GLM-4.5V were replaced by GLM-4.6V and GLM-4.6V-Flash (indicated by the double-headed arrow), keeping the total at 29.

| Model | Devel-oper | Reasoning mode | Japanese support | Engine | Engine version | Trans-formers | GPU | Temp | top_p | top_k | Other |
|---|---|---|---|---|---|---|---|---|---|---|---|
| Qwen3.5-397B-A17B | Alibaba | Both | Adopted | vLLM | 0.18.1 | 4.57.6 | GH200 ×12 | R: 0.6 / NR: 0.7 | R: 0.95 / NR: 0.8 | 20.0 | pres_pen=R: 0.0 / NR: 1.5 |
| Kimi-K2.6 | Moonshot | Both | Adopted | vLLM | 0.20.2rc1.dev65 | 5.8.0 | GH200 ×8 | R: 1.0 / NR: 0.6 | 0.95 | – | – |
| gemma-4-31B-it | Google | Both | Adopted | vLLM | 0.20.2rc1.dev65 | 5.8.0 | GH200 | 1.0 | 0.95 | 64.0 | – |
| Qwen3.5-122B-A10B | Alibaba | Both | Adopted | vLLM | 0.18.1 | 4.57.6 | GH200 ×4 | R: 1.0 / NR: 0.7 | R: 0.95 / NR: 0.8 | 20.0 | pres_pen=1.5 |
| Qwen3.6-35B-A3B | Alibaba | Both | Adopted | SGLang | 0.5.10rc0 | 5.3.0 | GH200 | R: 1.0 / NR: 0.7 | R: 0.95 / NR: 0.8 | 20.0 | pres_pen=1.5 |
| Qwen3.6-27B | Alibaba | Both | Adopted | vLLM | 0.20.2rc1.dev65 | 5.8.0 | GH200 | R: 1.0 / NR: 0.7 | R: 0.95 / NR: 0.8 | 20.0 | pres_pen=R: 0.0 / NR: 1.5 |
| gemma-4-26B-A4B-it | Google | Both | Adopted | vLLM | 0.20.2rc1.dev65 | 5.8.0 | GH200 | 1.0 | 0.95 | 64.0 | – |
| Qwen3.5-35B-A3B | Alibaba | Both | Adopted | SGLang | 0.5.10rc0 | 5.3.0 | GH200 | R: 1.0 / NR: 0.7 | R: 0.95 / NR: 0.8 | 20.0 | pres_pen=1.5 |
| Llama-4-Maverick-17B-128E-Instruct | Meta | Reasoning unsup-ported | Reference | vLLM | 0.20.2rc1.dev65 | 5.8.0 | GH200 ×8 | 0.6 | 0.9 | – | – |
| Qwen3.5-27B | Alibaba | Both | Adopted | SGLang | 0.5.10rc0 | 5.3.0 | GH200 | R: 1.0 / NR: 0.7 | R: 0.95 / NR: 0.8 | 20.0 | pres_pen=1.5 |
| Qwen3.5-9B | Alibaba | Both | Adopted | SGLang | 0.5.10rc0 | 5.3.0 | GH200 | R: 1.0 / NR: 0.7 | R: 0.95 / NR: 0.8 | 20.0 | pres_pen=1.5 |
| GLM-4.6V | Z.ai | Both | Reference | vLLM | 0.20.2rc1.dev65 | 5.8.0 | GH200 ×4 | 0.8 | 0.6 | 2.0 | rep_pen=1.1 |
| Llama-4-Scout-17B-16E-Instruct | Meta | Reasoning unsup-ported | Reference | vLLM | 0.18.1 | 4.57.6 | GH200 ×4 | 0.6 | 0.9 | – | – |
| EXAONE-4.5-33B | LGAI-EXAONE | Both | Adopted | vLLM* | 0.20.2rc1.dev65 | 5.3.0.dev0 | GH200 ×2 | 1.0 | 0.95 | – | pres_pen=1.5 |
| Qwen3.5-4B | Alibaba | Both | Adopted | SGLang | 0.5.10rc0 | 5.3.0 | GH200 | R: 1.0 / NR: 0.7 | R: 0.95 / NR: 0.8 | 20.0 | pres_pen=1.5 |
| InternVL3.5-14B | OpenGVLab | Both | Adopted | vLLM | 0.18.1 | 4.57.6 | GH200 | 0.6 | 0.95 | 50.0 | – |
| InternVL3.5-8B | OpenGVLab | Both | Adopted | vLLM | 0.18.1 | 4.57.6 | GH200 | 0.6 | 0.95 | 50.0 | – |
| GLM-4.6V-Flash | Z.ai | Both | Reference | SGLang | 0.5.10rc0 | 5.3.0 | GH200 | 0.8 | 0.6 | 2.0 | rep_pen=1.1 |
| Step3-VL-10B | StepFun | Reasoning only | Adopted | vLLM | 0.20.2rc1.dev65 | 5.8.0 | GH200 | 1.0 | 1.0 | 0.0 | – |
| MiniCPM-V-4.5 | OpenBMB | Both | Adopted | vLLM | 0.18.1 | 4.57.6 | GH200 | 0.6 | 0.95 | 20.0 | – |
| gemma-4-E4B-it | Google | Both | Adopted | vLLM | 0.20.2rc1.dev65 | 5.8.0 | GH200 | 1.0 | 0.95 | 64.0 | – |
| Nemotron-3-Nano-Omni-30B | NVIDIA | Both | Reference | vLLM | 0.20.2rc1.dev129 | 5.8.0 | RTX 6000 | R: 0.6 / NR: 0.2 | R: 0.95 / NR: 1.0 | R: / NR: 1.0 | – |
| Nemotron-Nano-12B-v2-VL | NVIDIA | Both | Reference | vLLM | 0.20.2rc1.dev129 | 5.8.0 | RTX 6000 | R: 0.6 / NR: 0.0 | R: 0.95 / NR: 1.0 | R: / NR: 1.0 | – |

*continued on next page*

| Model | Developer | Reasoning mode | Japanese support | Engine | Engine version | Transformers | GPU | Temp | top_p | top_k | Other |
|---|---|---|---|---|---|---|---|---|---|---|---|
| command-a-vision-112B | Cohere | Reasoning unsupported | Reference | vLLM | 0.20.2rc1.dev65 | 5.8.0 | GH200 ×6 | 0.3 | 0.75 | 0.0 | – |
| Phi-4-reasoning-vision-15B | Microsoft | Reasoning only | Reference | vLLM | 0.20.2rc1.dev129 | 4.57.6 | RTX 6000 | 0.8 | 0.95 | 50.0 | – |
| Molmo2-8B | AI2 | Reasoning unsupported | Reference | vLLM | 0.18.1 | 4.57.6 | GH200 | 0.0 | – | – | – |
| Phi-4-multimodal-instruct | Microsoft | Reasoning unsupported | Reference | vLLM | 0.20.2rc1.dev129 | 5.8.0 | RTX 6000 | 0.0 | 1.0 | 1.0 | – |
| aya-vision-8b | Cohere | Reasoning unsupported | Adopted | vLLM | 0.18.1 | 4.57.6 | GH200 | 0.3 | – | – | – |
| Molmo2-O-7B | AI2 | Reasoning unsupported | Reference | vLLM | 0.20.2rc1.dev65 | 5.8.0 | GH200 | 0.0 | 1.0 | – | – |

**Table S1:** Model characteristics and inference parameters. Reasoning mode: Both = reasoning enabled and disabled modes available; Reasoning only = model always reasons; Reasoning unsupported = reasoning mode not available. Japanese support: Adopted = Japanese explicitly supported or language unspecified in model card; Reference = model card lists supported languages but Japanese is not included. For models with different generation parameters for reasoning-enabled (R) and reasoning-disabled (NR) modes, values are shown as ”R: value / NR: value”; identical values are shown once. A dash indicates that the parameter was not set (engine default). GH200 = NVIDIA GH200 Grace Hopper; RTX 6000 = NVIDIA RTX 6000 Pro Blackwell Max-Q (96 GB VRAM); ×n after the GPU name indicates pipeline parallelism across n nodes. vLLM* = vllm-exaone45 fork. pres_pen = presence_penalty; rep_pen = repetition_penalty.

Supplementary Data. Per-model bootstrap effect sizes with confidence intervals and permutation test results, text-only accuracy, accuracy by examination year, and answer-extraction success rates by model and condition are available at https://github.com/miscelllaneous/jdrbe-vlm.
